# Esketamine rapid antidepression combined with dexmedetomidine sleep modulation for patients with depression and insomnia

**DOI:** 10.1101/2024.11.07.24316911

**Authors:** Muyan Zuo, Yaozu Li, John P Williams, Yongxiang Li, Lina Sun, Ruoguo Wang, Guoqiang Ren, Qinyan Xu, Jianxiong An

## Abstract

**Purpose:** Depression combined with insomnia is a complex bidirectional relationship that is more difficult to treat versus a single disorder, and there is a lack of effective treatments available. In this study, we carried out a novel way to simultaneously intervene in depression combined with insomnia and examined functional magnetic resonance imaging (fMRI) to help further elucidate the mechanisms.

**Patients and methods:** A total of 105 patients with depression and insomnia were included in this observational and prospective study. 17-item Hamilton Depression Scale (HAMD-17), Pittsburgh Sleep Quality Index (PSQI) were collected from medical records at baseline (T0), 24 hours (T1), 7 days (T2), 14 days (T3), 21 days (T4), one month (T5), two months (T6), three months (T7) follow-up. fMRI scans were performed at baseline and two hours after treatment.

**Results:** Compared with baseline, the symptoms of depression in T1-T7 were significantly reduced. At two hours after treatment, the left amygdala, the left hippocampus, the left superior temporal gyrus, the left anterior cingulate gyrus, and the left paracingulate gyrus showed a consistent reduction in spontaneous neural functional activity. In contrast, the right dorsolateral superior frontal gyrus, middle frontal gyrus, infraorbital frontal gyrus, middle orbital frontal gyrus and right caudate nucleus showed increased consistency of spontaneous neural function. No unanticipated safety issues were detected, and the rate of side effects was equivalent to those reported in RCTs.

**Conclusion:** Our findings support the efficacy and safety of esketamine combined with dexmedetomidine (Dex) in patients with depression and insomnia, which provides a new approach to clinical improvement of depression combined with insomnia.

## Introduction

Depression is the most common mental disorder worldwide, accounting for 17.3% of the global burden of mental illness and is characterized by a variety of symptoms such as depressive mood, anhedonia, somatization, cognitive disturbance, and sleep disorder.^1^ Sleep disorder is always the primary complaint and the first symptom of depressed patients, suicidal ideation, treatment response, relapse risk, and the intensity of depression are all correlated with the severity of insomnia.^2^ Numerous studies have demonstrated that insomnia and depression have a complex bidirectional link rather than a simple cause-and-effect one.^3,4^ Additionally, sleep disorders linked to depression are persistent and troublesome conditions that significantly impair patients’ social functioning and quality of life.^5,6^

Sleep disorder was traditionally seen as being simply affiliated with depression thus, insomnia was rarely the target of treatment because it was widely assumed that insomnia will resolve with treatment for depression.^7^ Currently however, sleep disorder is recognized as a distinct diagnostic category that can potentially contribute to episodes of depression, and enhancing sleep quality has been found to have a positive impact on the outcomes of depression.^8^ The main treatment modalities currently available include medication and cognitive behavioral therapy (CBT). Sedative antidepressants shorten sleep latency and improve sleep efficiency, however, there are issues such as drug dependence and tolerance, and approximately 30% of people diagnosed with major depression do not achieve remission, even after undergoing treatment with various antidepressant medications.^9^ Although CBT is a significant non-pharmacological treatment for insomnia and has been demonstrated to be useful in reducing depression, there are still limits due to a shortage of specialist psychiatrists and geographic distance from providers.^10^ Furthermore, both pharmacological and CBT are slow to work, especially as many depressed patients are at imminent risk of suicide.

In clinical practice, depressed patients with insomnia are prone to encounter more serious symptoms and difficulties in therapy.^11^ If only depression is treated, the core symptoms may be in remission, but persistent insomnia remains a common residual symptom in depressed patients, and if an intervention for insomnia occurs without concomitant treatment for depression, it is difficult to cure due to relapse and may contribute to unpleasant clinical outcomes.

Depression and insomnia share a common biological mechanism. Functional magnetic resonance imaging (fMRI) studies have found that 39 brain regions, including the amygdala, hippocampus, dorsolateral prefrontal cortex, anterior and posterior cingulate cortices, are associated with insomnia and depression.^12^ In addition, inflammatory markers such as IL-6 and TNF were elevated in both insomnia and depression patients,^13,14^ the combined action of cholinergic and monoaminergic neurons modulates sleep rhythms but are also one of the well-known pathophysiological mechanisms of depression.^9^ The substantial correlation between insomnia and depression is evident, but the exact interaction between them is still uncertain, which is also the reason for the lack of effective intervention methods in clinic. Due to these shortcomings in traditional treatments, clinicians and researchers are exploring novel ways to improve the treatment of depression combined with insomnia. The US Food and Drug Administration (FDA) and Europe approved esketamine for the treatment of patients with treatment-resistant depression (TRD) in 2019, a series of studies have demonstrated the efficacy and safety of esketamine.^15,16^ In our previous study, we developed Patient-Controlled Sleep (PCSL) as a therapeutic modality to treat chronic intractable insomnia by using dexmedetomidine (Dex).^17^ Given the above context, we initiated a pilot study to evaluate the effectiveness of the combination of esketamine and Dex in the treatment of depression combined with insomnia.

## Patients and Methods

### Participants and study design

This was a single arm study. The clinical trial has been approved by the Institutional Review Board (IRB) of the Shandong Second Medical University (wyfy-2023-ky-057) and registered on Chinese Clinical Trial Registry (ChiCTR2300070756), which was performed from April 2023 to December 2023 at our center. The study protocol followed the Declaration of Helsinki. All participants gave written informed consent for participation in the study and were fully aware of the risks associated with off-label use, informed consent for minor participants were obtained from legal guardian. Eligibility criteria for patients were as follows: (1) Met the diagnostic criteria for depression and insomnia according to the fifth edition of the Diagnostic and Statistical Manual of Mental Disorders. (2) The patient had right-hand dominance. Patients were excluded as follows: (1) Contraindication of Dex and esketamine: heart block, intracranial hypertension, hyperthyroidism and narrow-angle glaucoma. (2) Patients with other types of sleep disorders such as sleep apnea, restless legs syndrome, etc. All participants underwent a physical examination and laboratory screening, including electrocardiography, routine hematology and chemistry tests, sex hormone tests and thyroid functional measurement. In addition, all patients underwent free fMRI scans. The 17-item Hamilton Depression Scale (HAMD-17)^18^ were used to characterize depressive symptoms by clinicians (We have obtained the copyright license for the scale). The Pittsburgh Sleep Quality Index (PSQI) ^19^ was used to assess severity of insomnia.

### Study procedures

Prior to treatment, patients were instructed to fast for six hours, but they were permitted to consume clear liquids for a period of two hours. Sleep monitoring was performed with a polysomnography (PSG) device (iRem-A, iphysio, Hangzhou, China) equipped with six electroencephalograph (EEG) leads (F3, F4, C3, C4, O1, and O2), two electrooculogram (EOG) leads (M1 and M2), submentalis electromyogram (EMG), anterior tibialis EMG (right and left legs), respiratory effort (thoracic and abdominal impedance), electrocardiogram (ECG) and pulse oximetry. The PSG electrodes were applied to the participants an hour before therapy. The PSG recordings complied with American Academy of Sleep Medicine (AASM) recommendations.^20^ A certified registered nurse (CRN) inserted a dedicated intravenous cannula and started an infusion of 500 mL of saline solution. During the procedure, patients were monitored with ECG, non-invasive blood pressure (BP) monitoring, oxygen saturation (SpO_2_) and bispectral index (BIS). A solution was prepared by diluting 200 mcg of Dex (HumanwellHealthcare(Group)Co., Ltd. Hubei, China) with 0.9% normal saline to a volume of 50 mL in a syringe. The resulting drug concentration was maintained at 4 mcg/mL, a Constant Speed Syringe Pump (Hopefusion ™, SP5, Weifang, China) was utilized to administer the solution through the intravenous cannula. Dex titration was performed subsequent to the preceding protocol,^17^ the titration technique was established by a seasoned anesthesiologist (An JX), and the attending anesthesiologist (Li YX) performed and documented the operation and evaluation, the anesthesiologist operated the constant speed syringe pump, employing a basal rate of 60 mL/h (4 mcg/min). At the same time, a trained physician judged the sleep stage of the patients based on PSG, and the titration was stopped when the characteristic spindles and K-waves were seen and the patients entered the N2 stage, the dosage of Dex at this time was recorded as D1. Then esketamine 0.25 mg/kg diluted to 50 ml was infused over 40 minutes. A trained rater assessed side effects two hours after start of infusion. All patients underwent fMRI scanning two hours after stopping the infusion. The BP was kept within 30% of the initial level, phenylephrine was used to raise blood pressure and nitroglycerin was used to lower it if the reading deviated from the intended range; Anisodamine was given if the heart rate (HR) fell below 40 bpm; If the previous course of treatment didn’t work, atropine and isoproterenol were made accessible. If agitation occurred during the application of esketamine, midazolam was given for sedation, but needed to be excluded from the study. The anesthesiologist made the decision about additional medical care.

Following therapy, patients were excluded if they reported discomfort or exhibited side symptoms, such as agitation, bradycardia, or hypotension. Participants who were able to endure the treatment were identified as potential candidates to proceed with Self-controlled sleep. The patient was assisted by a specialized nurse in self-controlled sleep 30 minutes before bedtime, 1.5 times the dose of D1 was drawn by a syringe and injected into a sterile cotton ball, which was placed under the patient’s tongue for 10 minutes to allow the Dex to be fully absorbed, the cotton ball was removed, and the patient lied down to rest and fall asleep naturally. If the patient woke up and was unable to sleep at night, added 0.5 times D1 in the same way. In a sound-insulated chamber, specially trained nurses assessed HR and SpO_2_. After discharge from the hospital, the patient continued to apply self-controlled sleep every night in the manner described above and arranged for a trained medical professional to follow up and adjust the Dex dosage according to the sleep situation.

Participants were required to maintain their present psychiatric medication regimen consistent for four weeks before the first infusion and for three weeks after treatment. Patients received another esketamine antidepressant treatment when the patient experienced a relapse, relapse was considered the reappearance of symptoms from a continuing episode that had been symptomatically repressed.^21^ To evaluate the efficacy of esketamine use, when the patients’ HAMD-17 score decreased by 50% overall from the baseline evaluation, they were classified as responders.^22^ In addition, remission was defined as a HAMD-17 score of <7.^23^ The HAMD-17 and PSQI scores were gathered by telephone surveys and online questionnaires. Two physicians who were not participating in the clinical trial conducted follow-up interviews. Rating scales were administered at baseline (T0) and at seven-time intervals after infusion: 24 hours (T1), 7 days (T2), 14 days (T3), 21 days (T4), one month (T5), two months (T6), and three months (T7). All personal data was recorded on the Case Record Form (CRF) and was kept totally private for the sole purpose of study. Personal data will be kept by the members of the research team.

### Magnetic Resonance Imaging Acquisition

The imaging was conducted utilizing a 3.0-T Magnetic Resonance Imaging (MRI) scanner (Signa HDxt, GE Medical Systems, Waukesha, WI, USA) that had an eight-channel phase array head coil. Two earplugs that were the right size were utilized to limit scanner noise, and plastic foam pads were employed to lessen head movement. All individuals were instructed to stay motionless during the scanning procedure, keeping their eyes closed but not thinking or dozing off. If a participant displayed signs of discomfort, their scanning was stopped.

To begin, all participants underwent T2-weighted imaging (T2WI) to rule out the possibility of asymptomatic lesions.

The following parameters were used to obtain resting-state fMRI data: total volume=200, repetition time (TR)=2,000 ms, echo time (TE)=30 ms, flip angle=90°, slice thickness=4.0 ms, matrix=64×64, field of view (FOV)=240×240 ms, number of slices=32, the duration of the session was 400 seconds. Collecting three-dimensional high-resolution T1-weighted anatomical images with the following parameters: TR=7.8 ms, TE=3.0 ms, flip angle=15°, slice thickness=1.0 mm, FOV=256×256 mm^2^, matrix=256×256, and number of slices=188 were achieved using the defective gradient recalled acquisition, the entire session lasted 250 s.

### Data Preprocessing

REST plus, V1.25 was executed on the MATLAB 2017b platform (MathWorks, Natick, MA, USA), was used for processing fMRI data.^24^ The steps were as follow: (1) for magnetic field stabilization, delete the first ten of the 200-time points, (2) a correction to the slice timing in order to account for acquisition delays between slices, (3) head motion correction, (4) normalization, (5) remove the impact of thermal drift on the signal during extended scanning, (6) to remove the head motion confound, the 24 head motion characteristics were regressed out.^25^ As a result of excessive head motion, eleven participants were disqualified from additional analysis. (7) band-pass filtration with a 0.01–0.08 Hz frequency band.

### Regional homogeneity calculations

Regional homogeneity (ReHo) was evaluated using the Kendall’s coefficient of concordance (KCC), a task executed utilizing the REST plus toolkits version 1.25.^24^ In order to obtain the ReHo value, every 27 closest neighboring voxels’ time course KCC was calculated.^26^ For purposes of standardization, each voxel’s ReHo value was divided by the individual’s global mean ReHo. After the ReHo calculation, fullwidth at half-maximum [FWHM] = 6 mm was used for the spatial smoothing.

### Statistical analysis

The G*Power software and the ANOVA: repeated measures, within factors test were utilized for calculating the sample size. In accordance with previous research, the sample size was determined using the anticipated response to esketamine as 40%, a significance level of 0.05, and a power of 95%. Additionally, the non-experimental sample was accounted for when testing the hypothesis of 20% of the patients discontinuing treatment prematurely or not initiating it.

The mean and standard deviation (SD) are used to show continuous variables. Numbers and percentages are used to show category variables. Using a repeated-measures analysis of variance (ANOVA) on the HAMD-17 and PSQI scores, the intervention impact was determined. Pearson’s correlation analyses were used to investigate the associations between clinical symptom remission (lower HAMD-17 and PSQI scores) and sociodemographic characteristics.

A paired t-test was performed in the field of Data Processing & Analysis for Brain Imaging (REST plus, V1.25) ^24^ to compare the ReHo differences between the baseline and after treatment. In the statistical study, Gaussian Random Field theory (GRF) correction for multiple comparisons, the results were thresholded with voxel level *p*<0.005, cluster level *p*<0.05.

## Results

### Baseline characteristics and treatments

A total of 98 patients were enrolled, 41 of whom completed the second fMRI (Figure 1). Most participants completed the treatment phase and entered the post-treatment follow-up phase, with 77 completing the 3-month follow-up (Table 1). There were 47 (61.0%) females and 30 (39.0%) males, 30 (39.0%) married, and 47 (61.0%) singles. The mean age was 28.95 (±14.64) years, the mean duration of depression was 5.16 (±6.18) years, and the mean duration of insomnia was 4.98 (±6.82) years. The mean HAMD-17 score was 25.61 (±6.31) and the mean PSQI score was 14.08 (±2.84). 48 patients (62.3%) had a history of suicide ideation, and 18 patients (23.4%) had a history of self-injury.

**Figure 1.**
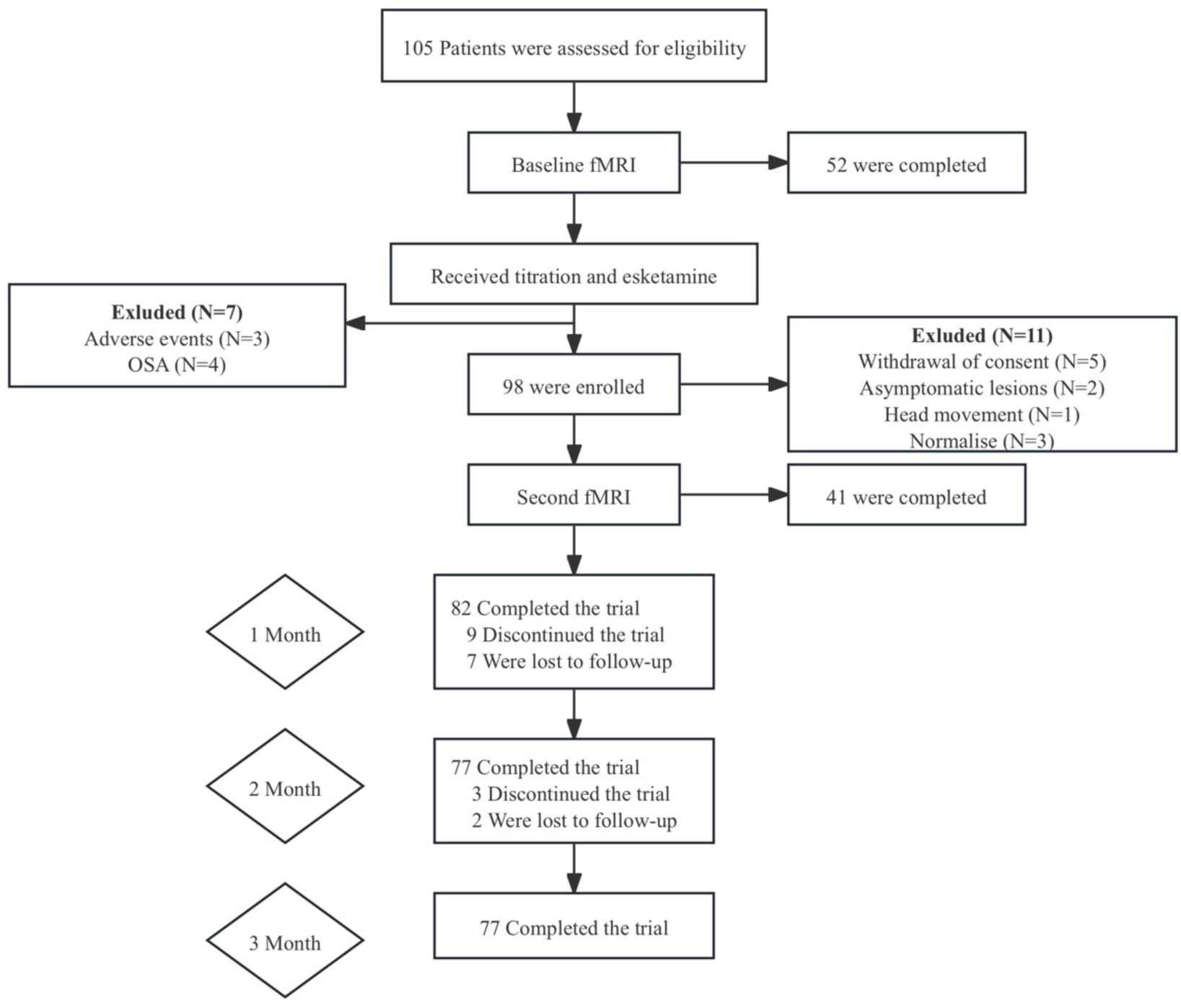
The flowchart for the study. **Abbreviations:** obstructive sleep apnea; fMRI: functional magnetic resonance imaging

**Table 1.** Sociodemographic and clinical data (n=77)

|  | Mean | Sd |
| --- | --- | --- |
| Age | 28.95 | 14.64 |
| Body mass index (calculated as kg/m<sup>2</sup>) | 23.75 | 5.34 |
| Education | 12.23 | 3.86 |
| Duration of depression (years) | 5.16 | 6.18 |
| Duration of insomnia (years) | 4.98 | 6.82 |
| Baseline clinical measures |  |  |
| HAMD-17 | 25.61 | 6.31 |
| PSQI | 14.08 | 2.84 |
|  | N | % |
| Female | 47 | 61.0% |
| Status |  |  |
| Single | 47 | 61.0% |
| Married | 30 | 39.0% |
| Previous suicidal ideation | 48 | 62.3% |
| Previous self-injury | 18 | 23.4% |
| Psychiatric comorbidities |  |  |
| BD | 10 | 13.0% |
| SZ | 8 | 10.4% |
| OCD | 3 | 3.9% |
| PTSD | 1 | 1.3% |
| SAD | 1 | 1.3% |
| Participants who attempted FDA-approved rTMS | 14 | 18.2% |
| Participants who attempted FDA-approved ECT | 13 | 16.9% |
| Add-on therapies |  |  |
| Hypnotics | 31 | 40.3% |
| SSRIs | 33 | 42.9% |
| SNRIs | 6 | 7.8% |
| Antipsychotic | 23 | 29.9% |
| Other antidepressants | 7 | 9.1% |

**Abbreviations:** HAMD-17: Hamilton Depression Scale; PSQI: Pittsburgh Sleep Quality
Index; BD: Bipolar disorder; SZ: Schizophrenia; OCD: Obsessive-compulsive disorder;
PTSD: Post-traumatic stress disorder; SAD: Social anxiety disorder; SSRI: Selective
serotonin reuptake inhibitors. SNRIs: Serotonin and Noradrenalin Reuptake Inhibitors.

Bipolar disorder (BD) was the most common comorbidity (13.0%), together with schizophrenia (SZ) (10.4%). 70.1% of the individuals had no additional mental conditions. 62 patients (81%) had received antidepressant therapy in the past, 14 patients (18.2%) had previously used repetitive transcranial magnetic stimulation (rTMS); and 13 patients (16.9%) had previously used ECT for treating depression. In addition, 31 patients (40.3%) were taking hypnotics. As part of augmentation techniques, 33 patients (42.9%) were taking SSRIs and 6 patients (7.8%) were using SNRIs as antidepressants, 23 patients (29.9%) were taking an antipsychotic in addition to antidepressant (Table 1).

### Effects of depression and insomnia

Repeated measurements showed that HAMD-17 scores decreased significantly at T1–T7 compared with T0. The average HAMD-17 score was 25.61 (±6.31) at T0, 19.77 (±6.85) at T1, and 11.90±6.44 at T7, One-way ANOVA results showed that HAMD-17 scores significantly decreased over time (F=29.043, *p<*0.01) (Figure 2A). In addition, esketamine also had a significant effect on reducing suicidal ideation, with 48 patients had suicidal ideation in the past three months before treatment and 42 patients no longer had suicidal ideation at T8; in addition, 18 patients had non-suicidal self-injury before treatment, and only two patients still had this behavior at T7.

**Figure 2.**
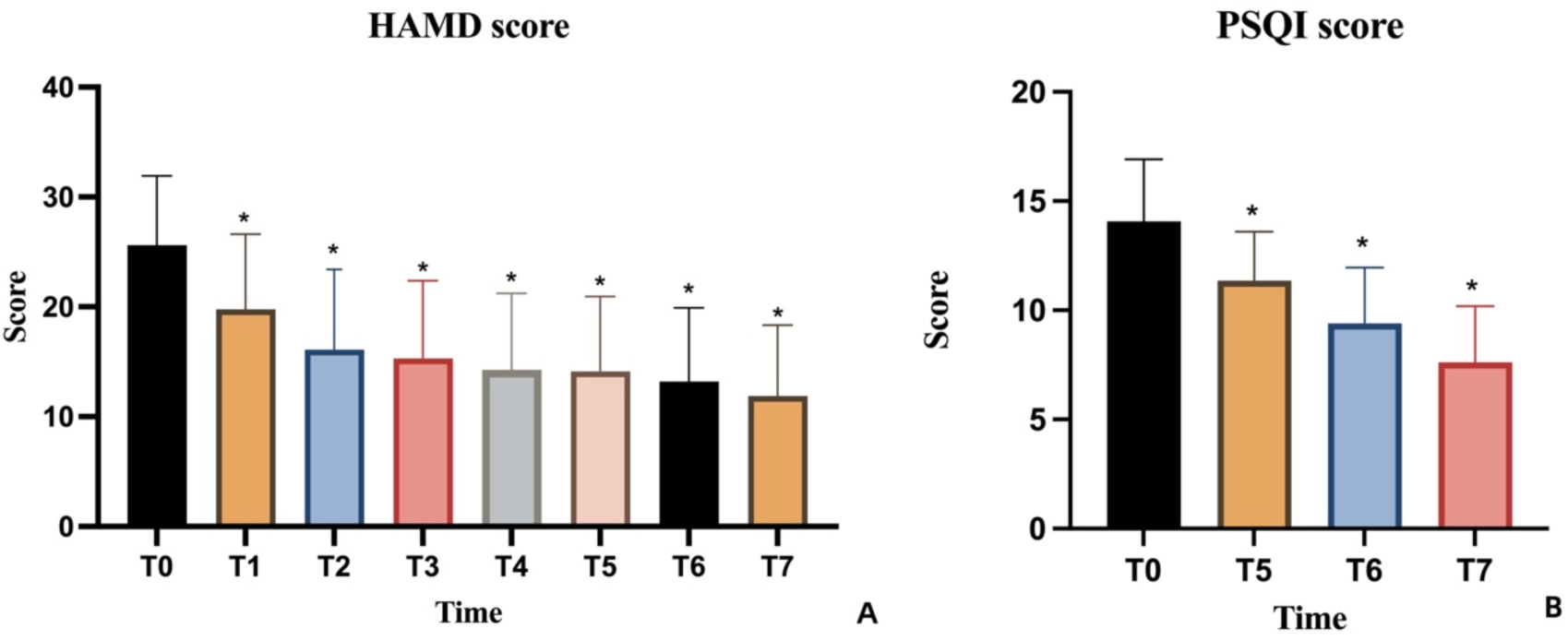
Treatment outcomes. **Abbreviations:** HAMD: Hamilton Depression Scale; PSQI: Pittsburgh Sleep Quality Index.

The average PSQI score was 14.08 (±2.84) at T0, 11.36 (±2.24) at T5 (*p<*0.01), and 9.40±2.56 at T6, and 7.61 (±2.58) at T7. PSQI scores decreased significantly at T5, T6 and T7 compared with T0 (F=107.852, *p<*0.01) (Figure 2B).

Overall effectiveness was assessed at T7, according to the definitions of responder and remission, 26 (34%) patients exhibited a clinical response to treatment, and 21 (27%) patients were in remission, 11(14%) patients did not respond to treatment and 19 (25%) patients had a HAMD-17 decrease of ≥20% but not yet 50% (Figure 3).

**Figure 3.**
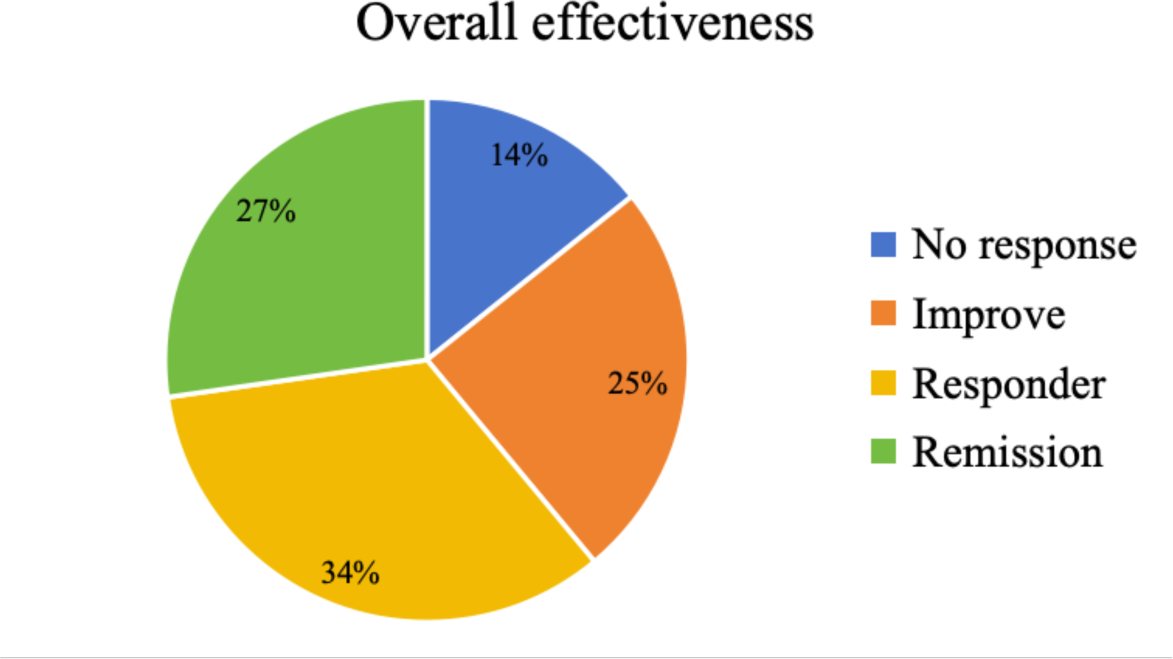
Overall effectiveness.

We further analyzed the demographic information and clinical data of responders and non-responders and found that there were statistically significant differences in age, insomnia duration, and marital status between the two groups (Table 2).

**Table 2.** Differences in sociodemographic and clinical data between responders and non-responders.

|  | responders<br>(n=47) | non-responders<br>(n=30) | P |
| --- | --- | --- | --- |
| Age | 24.60±13.13 | 35.77±14.46 | 0.001 |
| Body mass index (calculated as kg/m<sup>2</sup>) | 23.12±5.72 | 24.72±4.62 | 0.20 |
| Education | 11.91±3.99 | 12.73±3.67 | 0.37 |
| Duration of depression (years) | 4.13±5.42 | 6.79±6.99 | 0.06 |
| Duration of insomnia (years) | 3.49±5.34 | 7.30±8.22 | 0.02 |
| Baseline clinical measures |  |  |  |
| HAMD-17 | 25.40±6.62 | 25.93±5.89 | 0.72 |
| PSQI | 13.72±2.40 | 14.63±3.38 | 0.21 |

|  | N (%) |  | P |
| --- | --- | --- | --- |
| Female | 31 (66%) | 16 (53.3%) | 0.27 |
| Status |  |  |  |
| Single | 35 (74.5%) | 12 (25.5%) | 0.002 |
| Married | 12 (40.0%) | 18 (60.0%) |  |
| Previous suicidal ideation | 32 (68.1%) | 16 (53.3%) | 0.19 |
| Previous self-injury | 14 (29.8%) | 4 (13.3%) | 0.10 |
| Participants who attempted FDA-approved rTMS | 8 (17.0%) | 6 (20.0%) | 0.74 |
| Participants who attempted FDA-approved ECT | 6 (12.8%) | 7 (23.3%) | 0.23 |

### Brain activity between baseline and two hours after treatment

Compared with baseline, decreased ReHo values were observed in the left amygdala, left hippocampus, left superior temporal gyrus, left anterior cingulate gyrus, and paracingulate gyrus. In contrast, increased ReHo values were observed in the right dorsolateral superior frontal gyrus, middle frontal gyrus, inferior orbital frontal gyrus, middle orbital frontal gyrus, and right caudate nucleus (Figure 4). Table 3 details the regions with significant differences. The preceding outcomes were adjusted utilizing the Gaussian Random Field theory (GRF) (voxel *p*<0.005, cluster *p*<0.05).

**Figure 4.**
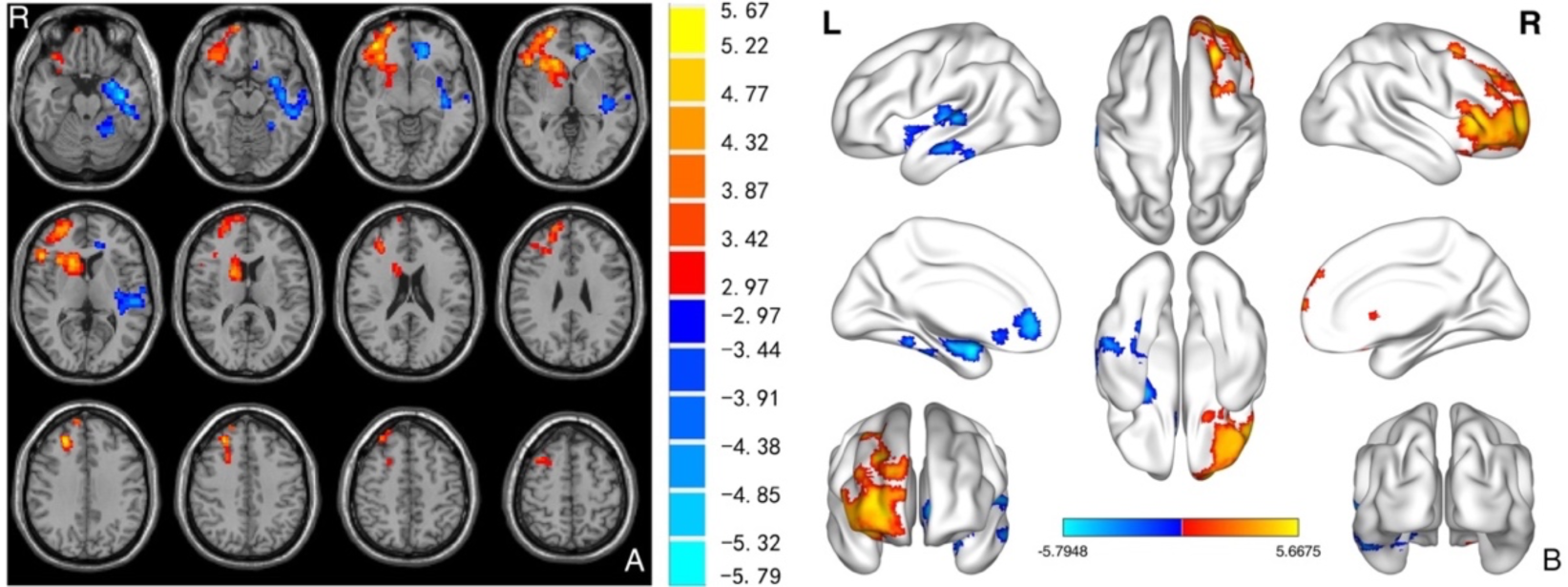
ReHo maps demonstrate significant variations in brain activity at baseline and after therapy.

**Table 3.**
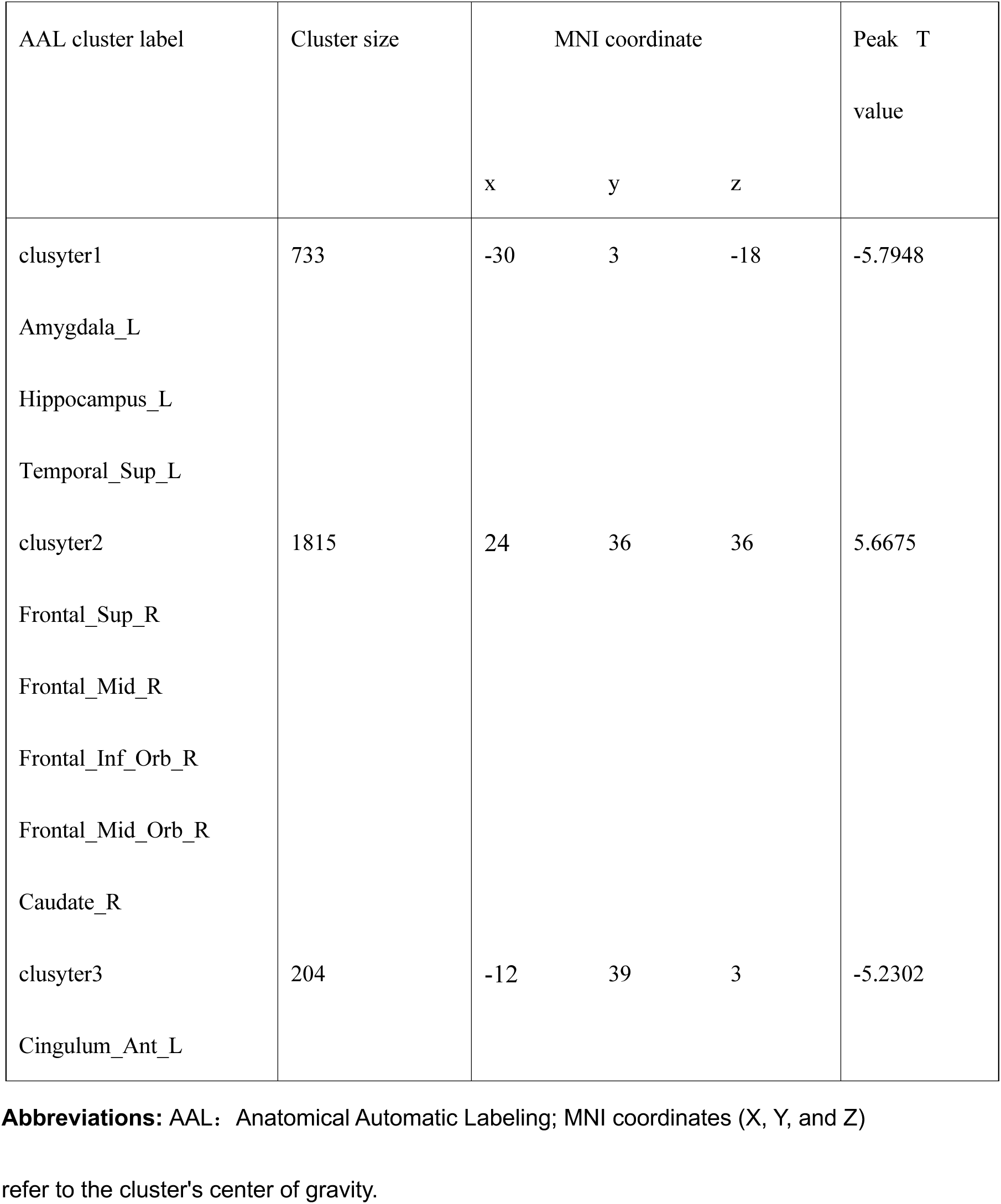
Brain areas with significantly changed ReHo values.

### Correlation analyses

There was no statistically significant correlation between baseline HAMD-17 score and age, and after treatment, percentage improvement in HAMD-17 scores was negatively correlated with age (*p*<0.001, r=0.4967) (Figure 5).

**Figure 5.**
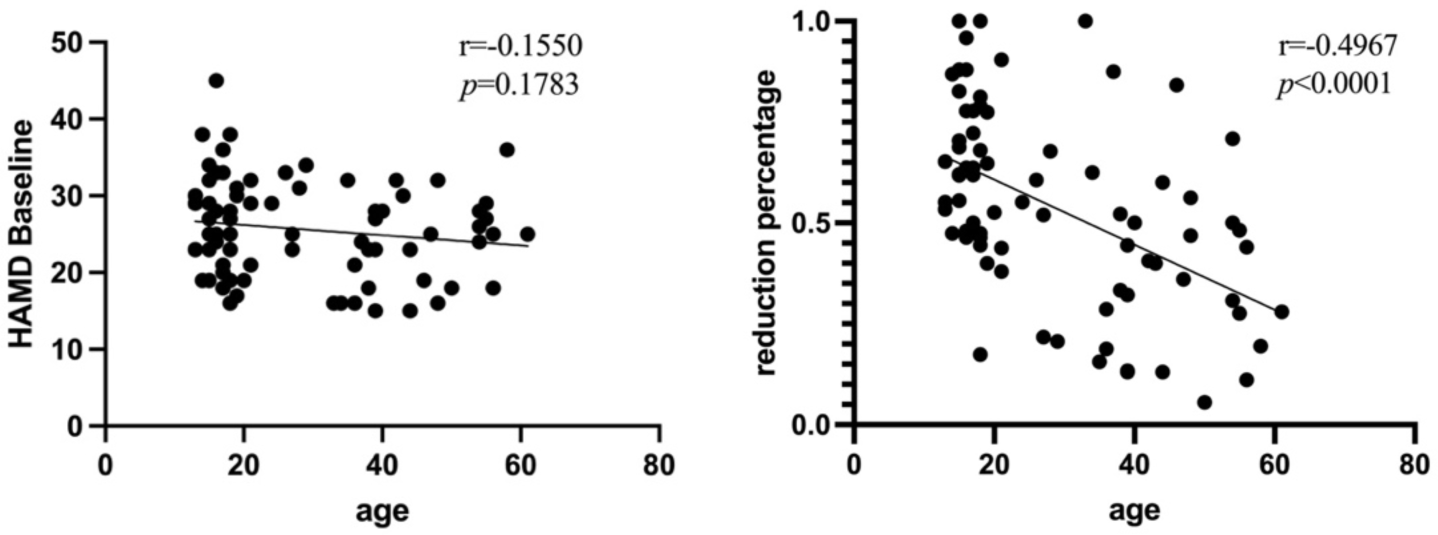
Correlation analyses between HAMD-17 scores and age. **Abbreviations:** HAMD: Hamilton Depression Scale.

### Safety and tolerability

As described above, seven patients discontinued treatment due to severe side effects. Three patients were diagnosed with OSA during titration, two patients had HR less than 40 after Dex titration, and one had severe psychomotor agitation. The most commonly reported adverse effects were dissociative symptoms (34.7%), dry mouth (30.6%), and vertigo (24.5%). Other adverse reactions included nausea, somnolence, increased blood pressure and headache (Figure 6).

**Figure 6.**
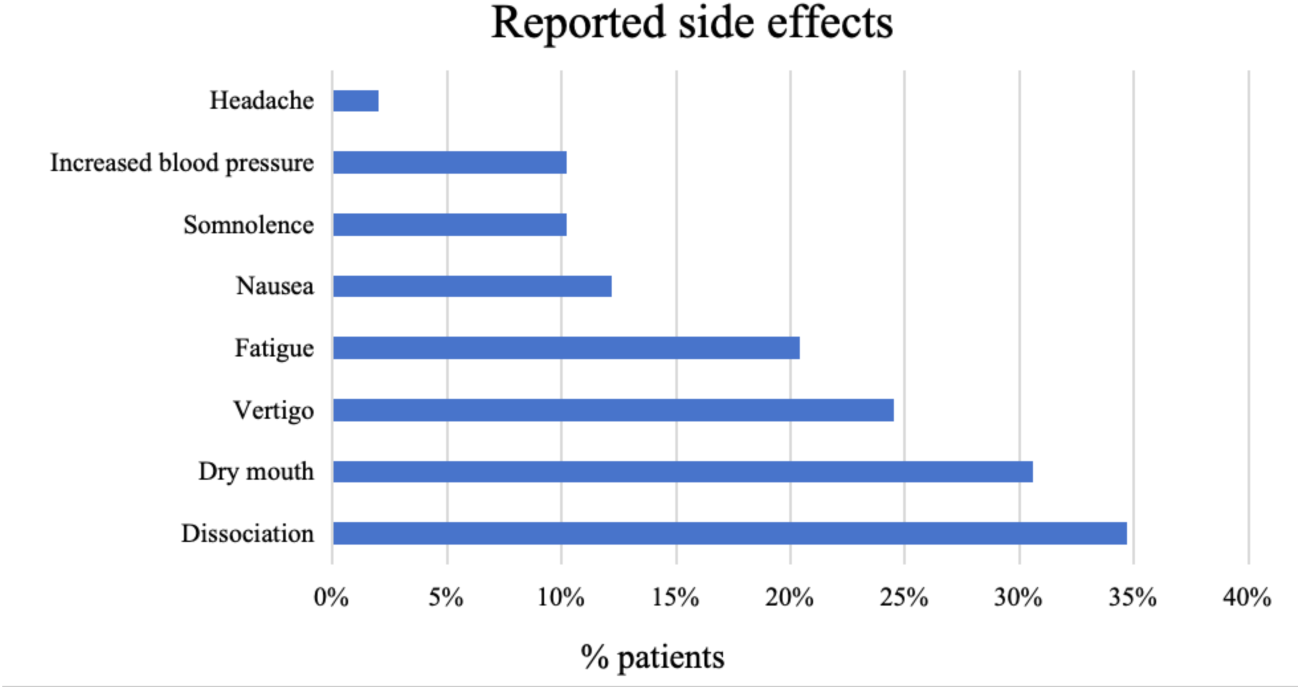
Reported side effects.

## Discussion

Through this study, we examined the efficacy, safety, and tolerability of esketamine in combination with Dex in the treatment of depression combined with insomnia. In the traditional way, approved oral combinations of antipsychotics and monoaminergic antidepressants have response rates of roughly 37%–56% after 6-12 weeks for TRD or insufficiently responsive MDD.^27^ In this study, based on HAMD-17 score and clinical symptom assessment, the response rate was 61%, as reported in a previous study, esketamine improved depressive symptoms more significantly,^28,29^ which is consistent with our findings. Importantly, we found that the improvement in depression was inversely associated with age, also, there was a difference in age between responders and non-responders, which is consistent with clinical observation. This age-related phenomenon was also found after esketamine application, and this decrease in efficacy may be related to increased neuromorphological alterations and treatment resistance, which may also increase the difficulty of treatment.

Dex is an alpha-2 agonist, frequently used in preoperative treatment, sedative, and antianxiety. Recent studies have found that Dex induces biomimetic sleep in humans, increases non-rapid eye movement (non-REM) time, and does not impair performance on psychomotor alertness tests compared to hypnotics.^30,31^

On the basis of PCSL, we made a further upgrade to the treatment, namely multimodal sleep (MMS);^32^ however, despite apparent success in treating patients with chronic intractable insomnia, there is still a proportion of insomnia patients with depression who relapse after treatment because of untreated depression. Thus, we developed a new set of approaches and procedures to address both issues simultaneously.

This treatment has several advantages over traditional treatments, first of all, recent study data demonstrated that Dex can considerably lower bipolar disorder patients’ two-hour agitation scores. These findings also suggested that Dex is crucial in lowering the high emotional risk of depression, manic episodes, and mixed mood states.^33^ Compared with prior randomized controlled trials (RCTs), our study included patients with psychiatric comorbidities and recent suicide risk, who might be considered more vulnerable to emotional states. Dex’s mood-stabilizing effects can be used as a useful adjuvant to esketamine antidepressants. Secondly, Dex can relieve sleep deprivation (SD) in rats by reducing oxidative stress of hippocampal neurons, restoring damaged cells, reducing hippocampal inflammation and improving cognitive function.^34^ Most depression patients suffer from accompanying cognitive disturbances such as memory decline and retardation of thought, so the long-term application of Dex may repair these symptoms. Not only that, we also found that there was a statistically significant difference in the duration of insomnia between responders and non-responders, and for patients with chronic insomnia, we believe that long-term repair with esketamine antidepressant combined with Dex sleep modulation that can improve the cognitive impairment caused by depression and insomnia. Finally, 62 patients (81%) received oral medications in the past in our study, and discontinuation and tapering were difficult steps for all patients who received oral antidepressants or sleeping pills as they were faced with discontinuation syndrome and relapse. Drugs that act on alpha2_C_ adrenoceptors may be useful in treating illnesses associated with increased startle responses and sensorimotor gating impairments, such as schizophrenia, drug withdrawal, attention deficit hyperactivity disorder, and posttraumatic stress disorder.^35^ As a highly selective alpha2 adrenergic receptor agonist, Dex may prevent withdrawal reactions in patients discontinuing the drug.

Due to the inconvenience in the application of patient-controlled device, we have adjusted the previous PCSL program. In this study, Dex was administered using a sublingual approach. The average onset of effect for the sublingual route was 13.89±1.54 minutes secondary to the abundant blood supply of the sublingual mucosa that permits direct and rapid absorption of the drug into the body circulation and avoids first-pass elimination. However, absorption appears to be influenced by a number of factors, such as the duration of contact between the drug and the mucosal surface, the local ph., and the salivary flow rate.^36^ In the future, better dosage forms and application methods may solve this problem.

In fMRI scans performed two hours after treatment, we observed significant changes. Elevated ReHo might be a sign of neuronal hyperactivity in a specific brain region.^26^ In fMRI studies of normal emotional processing, multiple brain regions are frequently co-activated when performing emotional tasks, possibly due to interactions between emotion production and regulation.^37^ Transcranial direct current stimulation (tDCS) treatment reduced depression and was linked to lower ReHo in the hippocampus and amygdala, which is consistent with our findings.^38^

The amygdala is a brain region that regulates emotions. People with depression exhibit elevated amygdala activation when exposed to favorable emotions. There is evidence that antidepressant medications or CBT can lower amygdala hyperactivity, which may be related to esketamine’s antidepressant effect.^39,40^ Furthermore, it was discovered that ReHo increased in the hippocampus of individuals with significant depression compared to healthy controls.^41^ Several investigations have demonstrated that hippocampus serves as a key node in the limbic thalamo-cortical network involved in cognitive functions as well as in the regulation of motivation and emotion.^42-44^ Changes in activity within the anterior cingulate cortex (ACC) have been associated with the antidepressant effects of ketamine. Similarly, changes in the ACC were observed in our study at an earlier time, two hours after injection. Esketamine may improve depression by modulating the overactive limbic system. In the future, nevertheless, more reliable experiments are required to test this theory.

A decrease in ReHo could potentially indicate an incoherent intraregional neural activity, while an increase in ReHo could indicate an improvement in emotional regulation. The superior frontal gyrus, dorsolateral (SFGdl) is involved in a variety of cognitive activities, primarily working memory, and the execution of cognitive maneuvers. The inferior frontal gyrus and middle frontal gyrus are essential components of the dorsolateral prefrontal cortex (DLPFC), which plays a role in the regulation of cognitive and emotional processes as well as the perception of pain.^45^

A study showed that compared to non-depressed patients, the brain of depressed patients typically has a reduced level of activity at rest in the region of the DLPFC,^46^ and ReHo in the DLPFC is elevated after rTMS.^47^ It has been noted that there was a significant increase in ReHo values in the right caudate nucleus after several courses of ECT in the previous study,^41^ and similar results were obtained in this study. We proposed an increase in ReHo after therapy could enhance emotional control or some cognitive functions to alleviate depressed symptoms.

Several limitations of our study deserve comment. First, the study’s open design and absence of a placebo or active comparison group are major shortcomings. Second, the duration of the study was relatively short, with induction and maintenance responses assessed for only three months. Depression is a chronic condition, and longer studies are necessary to fully determine whether the clinical benefits of esketamine can be maintained. Third, the study evaluated the efficacy and safety of the method but did not assess efficacy of two dosing regimens that differed in frequency of administration.

## Conclusion

This article is the first to propose that combining esketamine with Dex could be an effective treatment for patients with depression and insomnia. The safety and tolerability of esketamine combined with Dex for the treatment of depression and insomnia are supported by our observational data. But more research is required to determine long-term efficacy.

## Acknowledgements

Thanks to ShuangJie Cao MD. PhD., Kun Niu MD. PhD. from Institute for Innovation Diagnosis & Treatment in Anesthesiology, Shandong Second Medical University and Le Shi MD. PhD. from Peking University Sixth Hospital for their helpful suggestions.

## Ethics Approval and Consent to Participate

Ethical approval was given by the medical ethics committee of Shandong Second Medical University (number: wyfy-2023-ky-057). All participants gave written informed consent. Registration: Chinese Clinical Trial Registry (ChiCTR2300070756)

## CRediT authorship contribution statement

Muyan Zuo: collect clinical data, formal analysis and writing original draft. Yaozu Li: collect clinical data, formal analysis. John P Williams: modify the manuscript. Yongxiang Li, Lina Sun, Ruoguo Wang: carried out the study, collect clinical data. Guoqiang Ren: provided substantial comments. Qinyan Xu: fMRI scanning, data analysis. Jianxiong An: funding acquisition, design the study, writing - review & editing. All authors read and approved the final manuscript.

## Data availability statement

The data that support the findings of this study are available from the corresponding author upon reasonable request. Due to ethical restrictions, the data that support the findings of this study are not publicly available. Data may be available from the authors upon reasonable request and with the approval of relevant ethics committee.

